# Gendered pathways to adolescent mental health: An empirical assessment of a new conceptual framework

**DOI:** 10.64898/2026.06.09.26355310

**Authors:** Anita Alaze, Daniel Hagen, Tamara Schamberger, Oliver Razum, Céline Miani

**Affiliations:** Bielefeld University, School of Public Health, Department of Epidemiology and International Public Health, Bielefeld, Germany; Northwell Health, Department of Occupational Medicine, Epidemiology and Prevention, Great Neck, NY, USA; Donald and Barbara Zucker School of Medicine at Hofstra/Northwell, Department of Occupational Medicine, Epidemiology and Prevention, Hempstead, NY, USA; Donald and Barbara Zucker School of Medicine at Hofstra/Northwell, Department of Psychiatry, Hempstead, NY, USA; Feinstein Institutes of Medical Research, Institute of Behavioral Science, Manhasset, NY, USA; Bielefeld University, Faculty of Business Administration and Economics, Data Science Group, Bielefeld, Germany

**Keywords:** Structural Equation Modelling, Conceptual Framework, Gender, Adolescents, Mental health, Ethiopia

## Abstract

**Introduction:** Gender norms and roles are important determinants of physical and mental health in the key period of adolescence. Yet, the gendered pathways to mental health in adolescents are not fully understood. Using a conceptual framework for global adolescent mental health that we developed based on a Delphi process, we empirically investigated the associations between six gender-related constructs and adolescent mental health.

**Methods:** We used cross-sectional Gender and Adolescence: Global Evidence (GAGE) data from Ethiopia (2020) to explore the associations between sex, gender norms, psychological competencies, gender attitudes, gender roles, with the latter two also serving as mediators, and psychological distress (GHQ-12), using Structural Equation Modelling (SEM).

**Results:** The SEM model contained measurements from 1,584 adolescents, including 843 girls and 741 boys, with a median age of 13 years. Out of 14 pathways tested, we found statistically significant associations between psychological competencies and psychological distress; sex and gender attitudes; and between gender norms and psychological competencies, gender attitudes, and gender roles. Hence, the gender-related constructs were mostly associated with each other, rather than with psychological distress.

**Conclusion:** The gender-related constructs are strongly interrelated, thereby attenuating their individual effects on psychological distress. The interplay of gender-related constructs should be considered when developing interventions to promote mental health in adolescents.

## 1 Introduction

Adolescence is the first stage of life in which sex differences in mental health emerge in terms of morbidity and mortality (1). Mental health problems, such as depression, anxiety and suicidality, increase sharply during this period (2). This phenomenon and the magnitude of these problems have been reported globally (3). These sex differences are assumed to, at least in part, stem from gender norms, roles, and relations (4). However, it is not yet known through which specific pathways or mechanisms gender affects mental health outcomes in adolescents (5, 1).

Gender norms, roles, and relations are acquired through gender socialisation, defined as the “process by which individuals learn the cultural behaviours associated with the concepts of femininity or masculinity” (6). This process begins primarily in the *family* as parents directly or indirectly influence children to conform to expected gender norms, roles and relations (7). The effects of gender socialisation become particularly evident during adolescence (8).

The *school environment* can reinforce the gender socialisation process through teachers (e.g. gendered role assignments) and peers (e.g., bullying due to gender non-conformity) (9). Gender norms either incentivise or disincentivise specific health behaviours, potentially leading to adverse (mental) health outcomes (10). “Beliefs about appropriate roles for males and females regarding the division of paid labor, homework, and childcare” (11) are often referred to as gender role attitudes, although great variability in the conceptualisation of gender roles exists. Masculine gender role characteristics are a protective factor against depression and depressive symptoms (4). The use of *social media* can be associated with increased mental distress (12). Media and social media use play an important role in reproducing social pressures and increase the exposure to stereotyping, objectifying and sexualising representations, which in turn strengthen beliefs in, e.g., gender stereotypes and body ideals (13). Psychological *competencies*, such as coping skills, have been identified as strong determinants of well-being not only in individuals with mental health disorders but also in the absence of mental disorders (14).

To explore more complex gendered pathways to adolescent mental health, we previously developed a conceptual framework (hereafter referred to as the ‘GAM framework’) in an international Delphi study (15). It encompasses six gender-related constructs, namely sex assigned at birth, gender identity, gender norms (of the social environment), gender attitudes, gender roles, psychological competencies, and mental health. It also incorporates a range of socio-demographic variables that together form an intersectional lens. The gender-related constructs are hypothesised to be associated with adolescent mental health but have not yet been empirically tested. Therefore, we investigate multiple (mediated) regression models via Structural Equation Modelling (SEM) for analysis of the interlinked effects. SEM is considered an innovative approach for analysing sex/gender (16), is compatible with an intersectional approach (17), and is used in cross-sectional studies (18). We also conduct Exploratory (EFA) and Confirmatory Factor Analysis (CFA) to validate a newly developed gender norms scale for the dataset employed in this study. The EFA serves to explore and refine the scale’s underlying factor structure, while the CFA aims to confirm the hypothesised factor model of the EFA in a confirmatory setting (19).

## 2 Materials and Methods

### 2.1 Ethical approval

Our study did not require ethical approval, as it is based on the secondary analysis of previously collected, publicly available, fully anonymous, and non-identifiable third-party data. This data is available free of charge for any researcher following registration with the data provider ‘UK Data Service’. We have complied with the data provider’s terms of use.

### 2.2 Data

Our study draws on quantitative data from the Gender & Adolescence: Global Evidence (GAGE) project, a ten-year (2015-2024) mixed-methods longitudinal research and evaluation programme. GAGE surveys 18,000 adolescents aged 10 to 19 years and other community members across six countries in the Global South, with a particular focus on vulnerable adolescents, such as those with disabilities, who married before the age of 18, refugees, adolescent mothers and out-of-school youth (20). We use data from the second round of data collection in Ethiopia in 2020, as it includes validated tools for mental health and gender-related variables. The GAGE sample was collected purposefully (20).

### 2.3 Measures

We selected and operationalised measures guided by the GAM framework (15). Figure 1 is an adaptation of this framework based on data available in the Ethiopian dataset.

**Fig 1:**
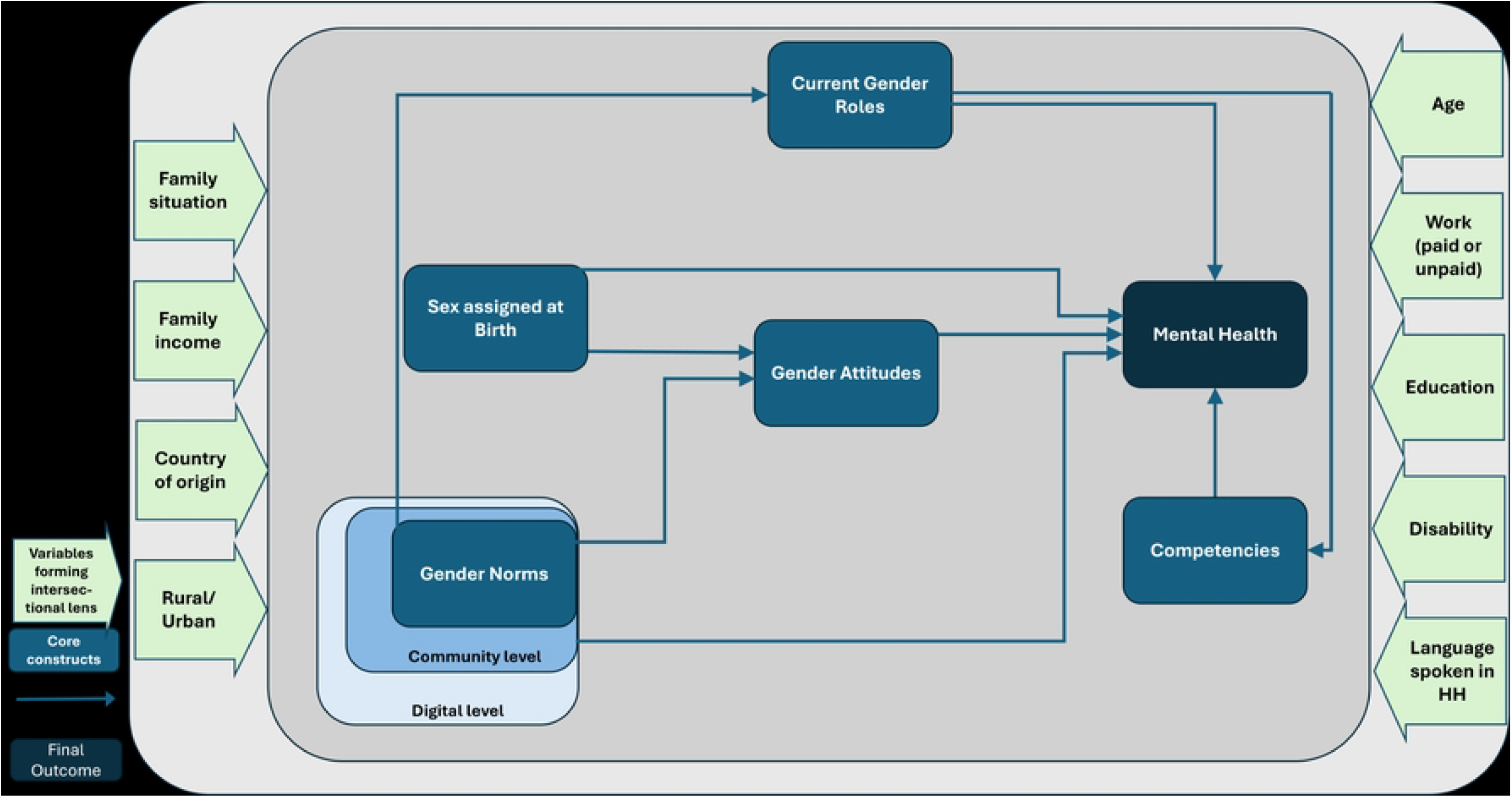
Adapted conceptual framework from a previous Delphi study (15) illustrating the operationalised constructs with the Ethiopian GAGE dataset.

We used the binary variable *sex* for the construct **sex assigned at birth**. We could not operationalise the construct of **gender identity** as no such measure exists in the dataset. While we could not operationalise the household and political levels of **gender norms** due to a lack of appropriate items, we operationalised the community level with a *peer violence index, being a club member*, and *having a role model*, and the digital level with *media* and *social media use*. We recoded these variables so that higher values indicate higher exposure to gender norms. A detailed description of the recoded variables is shown in Supplementary Information A.

We operationalised **gender attitudes** using the GAGE gender norms scale. In previous work (21), we explored the factor structure of this newly developed scale using data from an earlier GAGE round. The scale combines items from existing adolescent gender norms scales with items developed specifically for the GAGE project, but neither the original scales nor the GAGE scale have undergone prior empirical validation for the specific study population under investigation. Therefore, we explored its factor structure in the present sample. Following methodological recommendations and our previous approach, we used EFA, as the underlying factor structure could not be assumed a priori, and subsequently tested the hypothesised factor model of the EFA in a confirmatory setting using CFA (see Supplementary Information B) (19). Ultimately, we retained 12 of the original 32 items of the GAGE gender norms scale. We recoded the scale so that a higher score indicates more gender egalitarian attitudes. The revised scale demonstrated good internal consistency reliability (Cronbach’s α = 0.81).

**Gender roles** can be defined by the sexual division of labour, where specific tasks are assigned based on gender. Accordingly, we operationalised the gender roles construct with a *gendered activities score*. A higher score translates to more hours spent on activities per day, indicating adherence to more traditional gender roles.

We operationalised the construct **psychological competencies** with six instruments: a *voice* index; a r*esilience* index; an *index for talking to caregivers about work, religion, education, and bullying*; *speaking up in class*; *expression of opinion to friends;* and *to elders*. These variables were recoded so that higher values indicate a higher level of psychological competencies.

We operationalised the **mental health** construct using the validated instrument General Health Questionnaire (GHQ)-12. The GHQ-12 is a short and self-administered screening tool to detect symptoms of psychological distress and short-term changes in mental health (22). A higher GHQ-12 score translates to increased psychological distress.

### 2.4 Statistical Analysis

#### 2.4.1 Structural Equation Modelling

We employed SEM to simultaneously explore the hypothesised associations between the gender-related construct and adolescent mental health using a mix of measured, observed variables and unmeasured, latent variables. SEM subsumes various other statistical techniques, including CFA, path analysis, and multiple regression analysis.

We tested for multivariate normality (Shapiro-Wilk normality test), multicollinearity (Variance Inflation Factor), and common method bias (Herman’s Single Factor). We then developed and tested the *measurement model* for the latent variables using CFA. We used the Chi-squared test statistics, the Bentler Comparative Fit Index (CFI), the Tucker-Lewis Index (TLI), the Goodness-of-Fit Index (GFI), the Adjusted Goodness-of-Fit Index (AGFI), the Root Mean Square Error of Approximation (RMSEA), and the Standard Root Mean Square Residual (SMSR) to assess model fit (19, 23). Thresholds for good model fits are: CFI, TLI, GFI, AGFI ≥ 0.90, RMSEA < 0.06, and SMSR < 0.08 (19, 23). We assessed localised areas of model misfit using standardised factor loadings. While a threshold of 0.4 is commonly recommended, four indicators with loadings above 0.2 were retained because of their theoretical relevance to the GAM framework and their contribution to model fit. Consistent with recommendations to balance statistical and theoretical considerations in model evaluation, the final specification reflects a theory-informed compromise between indicator strength and model fit. We estimated parameters using maximum likelihood with robust standard errors to account for non-normality. We applied sample weights to ensure representativeness of the Ethiopian adolescent population, and we treated ordinal variables with ≥5 response categories as continuous. Subsequently, we specified a *structural model* to assess the extent to which the exogenous and mediator variables explain the variance in the dependent variables psychological competencies and the outcome mental health. This resulted in five regression and four mediation models. We obtained the share of mediation as the quotient of the absolute indirect effect by the absolute total effect. Following the intersectional lens in the GAM framework, we used a range of covariates as predictors for the two exogenous variables gender norms and sex. Statistical significance was assessed at the 5% level. We conducted all analyses in R version 4.4.1 (R Core Team, 2025). We estimated the CFA and SEM using the “lavaan” package version 0.6-19 (24), and we followed reporting guidelines for SEM (25).

## 3 Results

### 3.1 Sample Characteristics

Our sample includes measures of 1,584 adolescents with complete data. The descriptive statistics are presented in Table 3. The sample comprises a higher number of girls (53%) than boys (47%), with an average age of 13 years (standard deviation (SD): 0.9). The average GHQ-12 score is 7.1 (SD: 3.8), which is below the validated GHQ-12 cut-off of 9 for boys and 10 for girls for psychological distress and a potential mental disorder (26). The study participants reached a relatively high resilience score of 20 (SD: 4.0). The sample has slightly more equal than unequal gender attitudes with a value of 15 (SD: 5.6) and spends on average two hours per day on gendered activities (SD: 2.2). More adolescents express their opinions to their friends (78%) than to older people (45%). They talk on average about two of the four sensitive topics with their parents (SD: 1.2) and scored 19 on perceived influence in family matters. About half of the participants report having a role model (55%) and being a member of a club (46%). 87% and 39% of adolescents report not using social media and media, respectively, in the last 30 days. The peer violence index indicates low peer violence with 0.9 (SD: 1.8).

### 3.2 Structural Equation Modelling

The fully specified SEM model included data for 1,584 adolescents from 1,584 households. We found no multicollinearity as all variance inflation factors (VIF) were below 1.7 (threshold <5). Additionally, no common method bias was detected, with a proportion of variance of 0.22 (threshold <0.5). The SEM model simultaneously fitted latent variables, multiple regression models, and multiple mediation paths (Figure 2).

**Fig 2:**
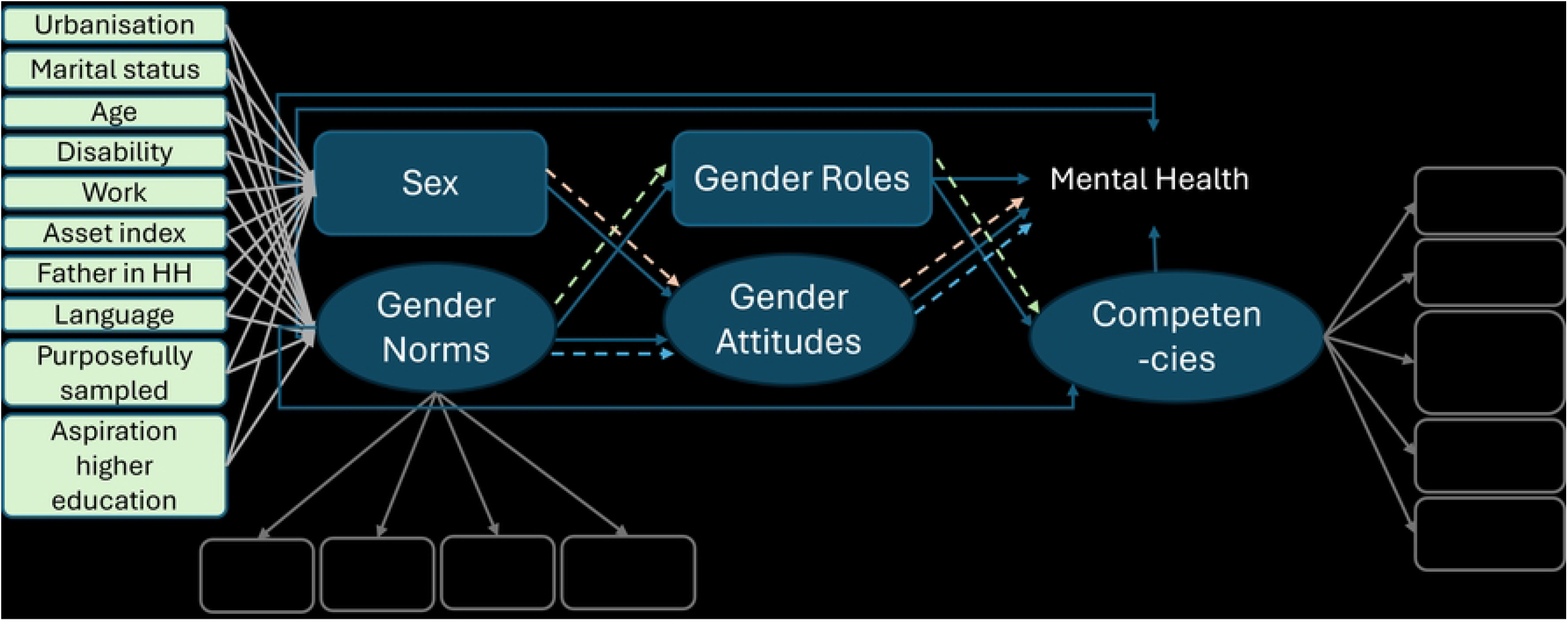
Illustration of a structural equation model examining how the gendered pathways (sex, gender norms, gender attitudes, gender roles, and competencies) influence adolescent mental health, with a set of covariates regressed on the two exogenous variables sex and gender norms. For simplicity reasons, we have omitted the error terms in the figure. Light green box = covariates and variables forming an intersectional lens Blue box = construct; Black box = final outcome; Grey box = indicator for the latent variable Oval box = latent variable (measured through observed indicator variables); Rectangular box = indicator for observed variable Full line arrow (blue) = regression path Dashed arrow (multiple colours) = mediation paths (one colour per mediation).

The fit statistics of the SEM model presented a mixed picture: The RMSEA and the SRMR (≤0.05) indicated a good absolute model fit and a good fit of the residuals. The CFI, TLI, GFI and AGFI with values of 0.76, 0.72, 0.89, and 0.82, respectively, fell below the recommended ≥0.90 threshold. The Chi-Squared test, with χ^2^(344) 1426.456, p<0.001, rejected the null hypothesis, which is common for models with sample sizes of over 400 observations (19).

Half of the analysed regression paths reached statistical significance (Table 2). Psychological competencies were negatively associated with psychological distress, which means that a higher level of competencies is, on average, associated with lower values of psychological distress and thus better mental health. Higher exposure to gender norms and more hours spent on gendered activities were, on average, associated with higher levels of competence, albeit only significantly for gender norms. Being a girl compared to being a boy and being more exposed to gender norms were positively and significantly associated with more egalitarian gender attitudes. Higher exposure to gender norms was negatively associated with more egalitarian gender roles, which means that higher exposure to gender norms is, on average, associated with fewer hours spent on gendered activities.

**Table 1:**
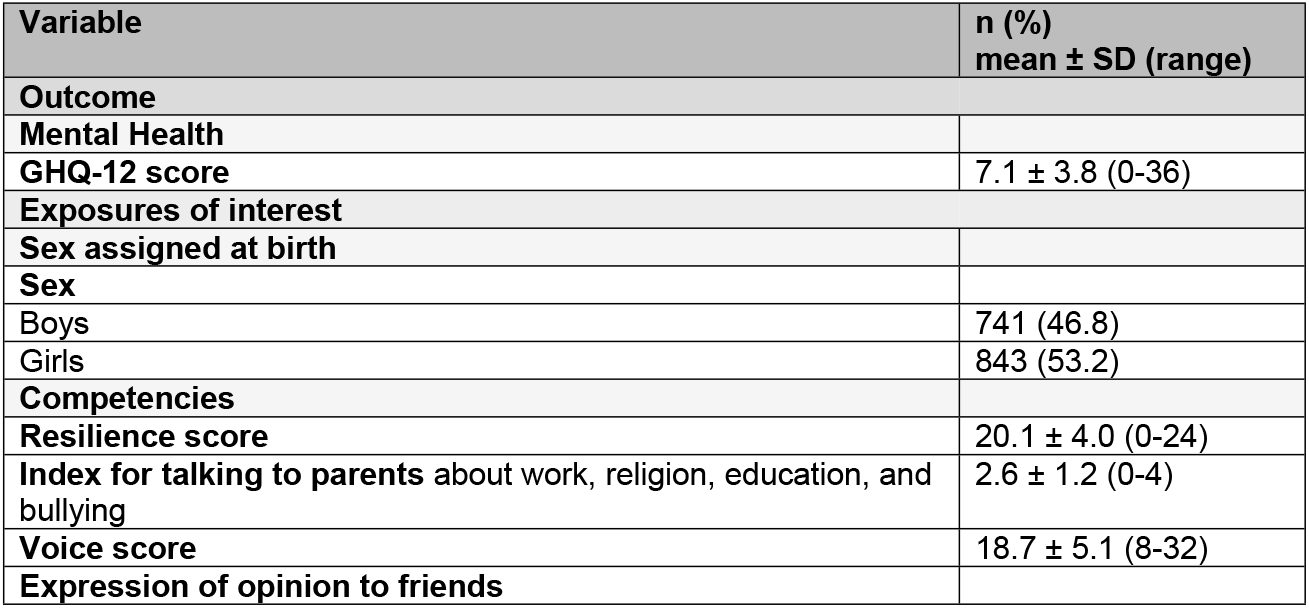
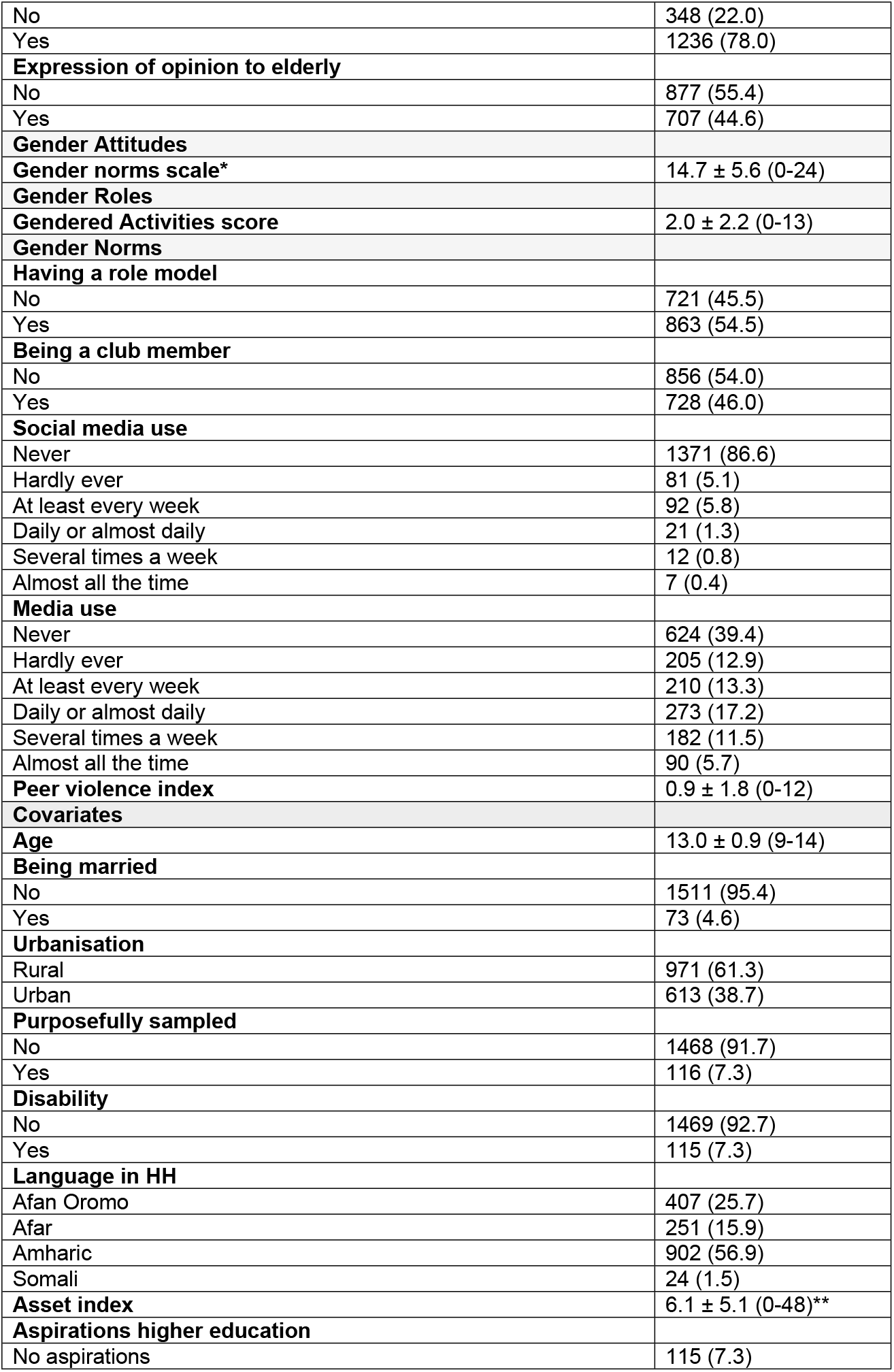
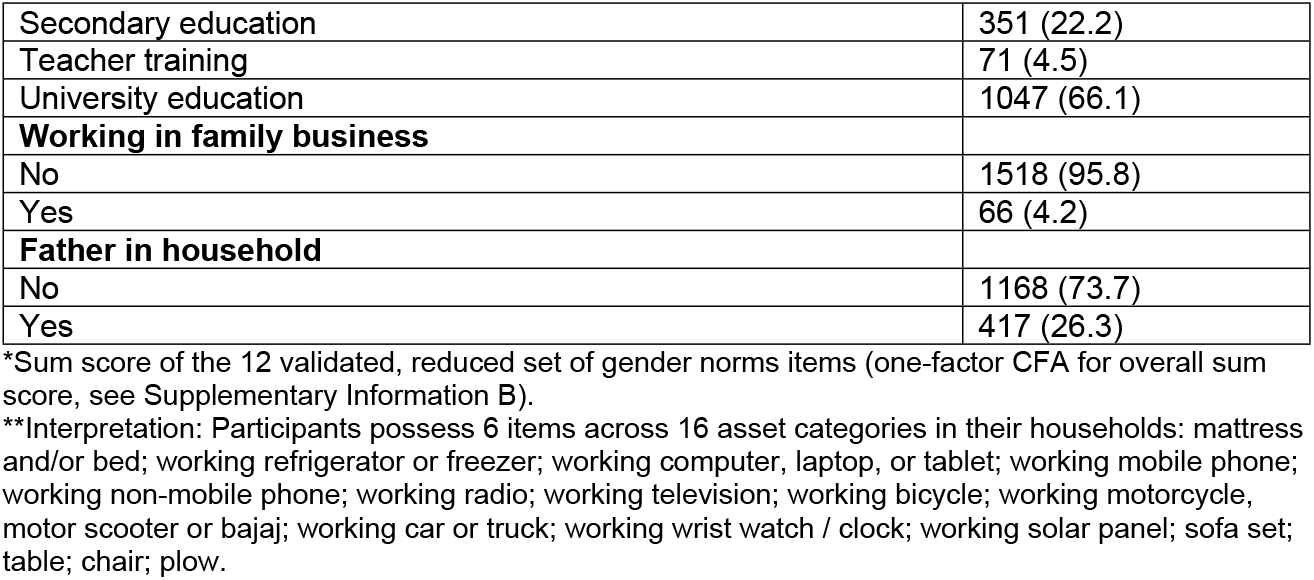
Descriptive statistics of the variables operationalising the GAM framework (n=1,584)

| Variable | n (%)<br>mean ± SD (range) |
| --- | --- |
| **Outcome** |  |
| **Mental Health** |  |
| **GHQ-12 score** | 7.1 ± 3.8 (0-36) |
| **Exposures of interest** |  |
| **Sex assigned at birth** |  |
| **Sex** |  |
| Boys | 741 (46.8) |
| Girls | 843 (53.2) |
| **Competencies** |  |
| **Resilience score** | 20.1 ± 4.0 (0-24) |
| **Index for talking to parents** about work, religion, education, and bullying | 2.6 ± 1.2 (0-4) |
| **Voice score** | 18.7 ± 5.1 (8-32) |
| **Expression of opinion to friends** |  |
| No | 348 (22.0) |
| Yes | 1236 (78.0) |
| **Expression of opinion to elderly** |  |
| No | 877 (55.4) |
| Yes | 707 (44.6) |
| **Gender Attitudes** |  |
| **Gender norms scale\*** | 14.7 ± 5.6 (0-24) |
| **Gender Roles** |  |
| **Gendered Activities score** | 2.0 ± 2.2 (0-13) |
| **Gender Norms** |  |
| **Having a role model** |  |
| No | 721 (45.5) |
| Yes | 863 (54.5) |
| **Being a club member** |  |
| No | 856 (54.0) |
| Yes | 728 (46.0) |
| **Social media use** |  |
| Never | 1371 (86.6) |
| Hardly ever | 81 (5.1) |
| At least every week | 92 (5.8) |
| Daily or almost daily | 21 (1.3) |
| Several times a week | 12 (0.8) |
| Almost all the time | 7 (0.4) |
| **Media use** |  |
| Never | 624 (39.4) |
| Hardly ever | 205 (12.9) |
| At least every week | 210 (13.3) |
| Daily or almost daily | 273 (17.2) |
| Several times a week | 182 (11.5) |
| Almost all the time | 90 (5.7) |
| **Peer violence index** | 0.9 ± 1.8 (0-12) |
| **Covariates** |  |
| **Age** | 13.0 ± 0.9 (9-14) |
| **Being married** |  |
| No | 1511 (95.4) |
| Yes | 73 (4.6) |
| **Urbanisation** |  |
| Rural | 971 (61.3) |
| Urban | 613 (38.7) |
| **Purposefully sampled** |  |
| No | 1468 (91.7) |
| Yes | 116 (7.3) |
| **Disability** |  |
| No | 1469 (92.7) |
| Yes | 115 (7.3) |
| **Language in HH** |  |
| Afan Oromo | 407 (25.7) |
| Afar | 251 (15.9) |
| Amharic | 902 (56.9) |
| Somali | 24 (1.5) |
| **Asset index** | 6.1 ± 5.1 (0-48)** |
| **Aspirations higher education** |  |
| No aspirations | 115 (7.3) |
| Secondary education | 351 (22.2) |
| Teacher training | 71 (4.5) |
| University education | 1047 (66.1) |
| **Working in family business** |  |
| No | 1518 (95.8) |
| Yes | 66 (4.2) |
| **Father in household** |  |
| No | 1168 (73.7) |
| Yes | 417 (26.3) |

\*Sum score of the 12 validated, reduced set of gender norms items (one-factor CFA for overall sum score, see Supplementary Information B).
\*\*Interpretation: Participants possess 6 items across 16 asset categories in their households: mattress and/or bed; working refrigerator or freezer; working computer, laptop, or tablet; working mobile phone; working non-mobile phone; working radio; working television; working bicycle; working motorcycle, motor scooter or bajaj; working car or truck; working wrist watch / clock; working solar panel; sofa set; table; chair; plow.

**Table 2:**
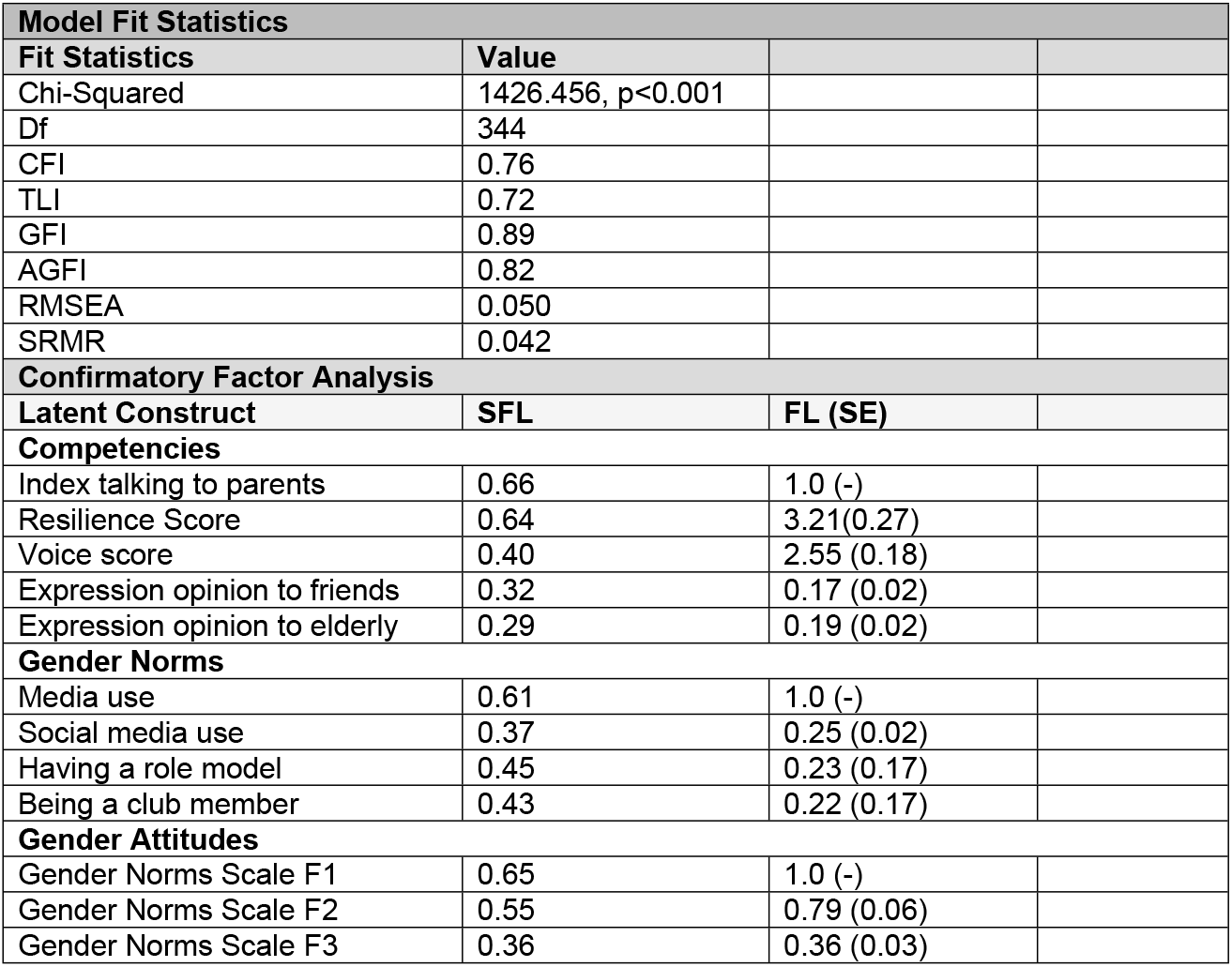
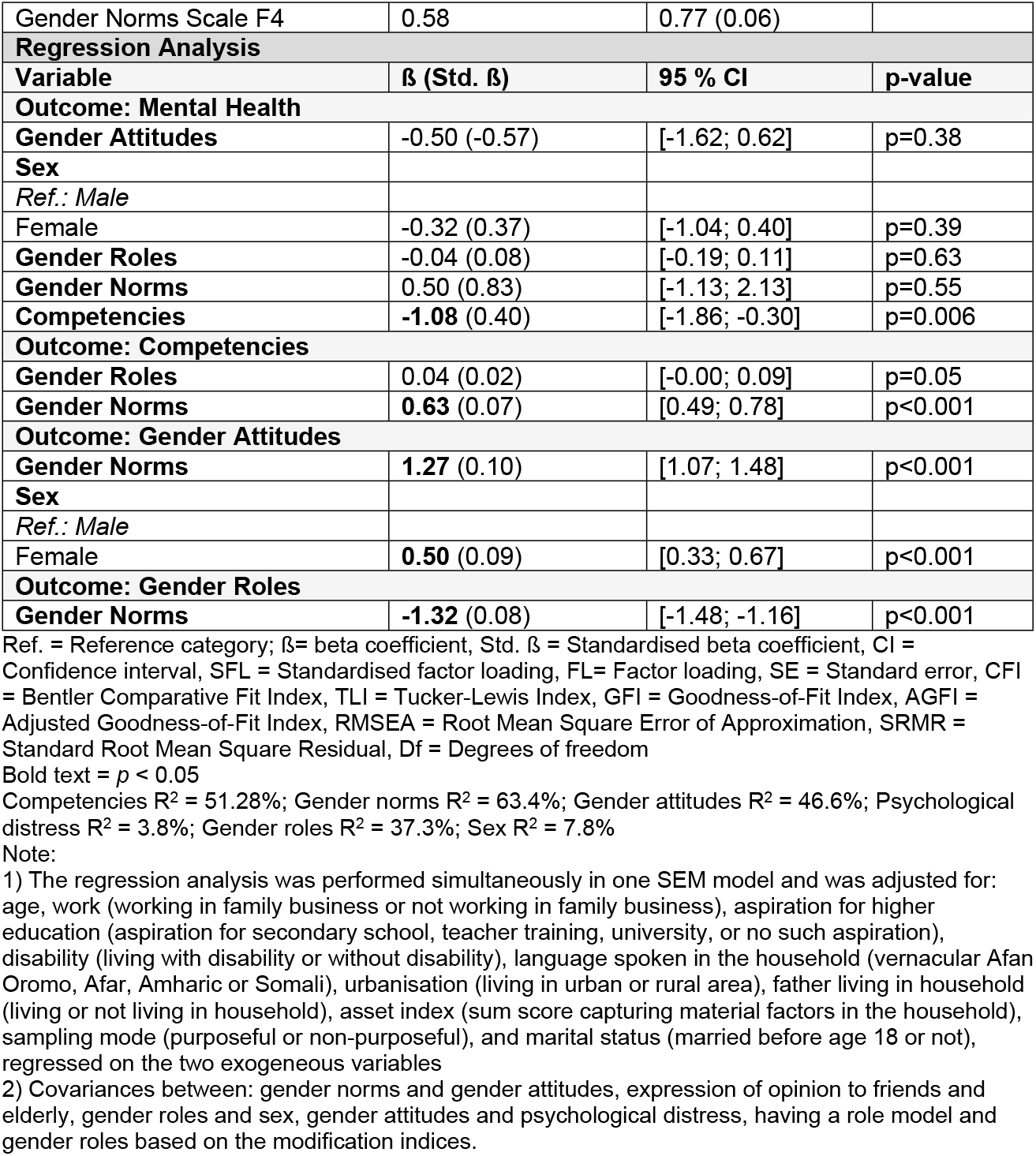
Results of the model fit, the latent variables, and the regression analysis in the SEM model (N=1,584)

| **Model Fit Statistics** |  |  |  |
| --- | --- | --- | --- |
| **Fit Statistics** | **Value** |  |  |
| Chi-Squared | 1426.456, $p < 0.001$ | | |
| Df | 344 |  |  |
| CFI | 0.76 |  |  |
| TLI | 0.72 |  |  |
| GFI | 0.89 |  |  |
| AGFI | 0.82 |  |  |
| RMSEA | 0.050 |  |  |
| SRMR | 0.042 |  |  |
| **Confirmatory Factor Analysis** |  |  |  |
| **Latent Construct** | **SFL** | **FL (SE)** |  |
| **Competencies** |  |  |  |
| Index talking to parents | 0.66 | 1.0 (-) |  |
| Resilience Score | 0.64 | 3.21(0.27) |  |
| Voice score | 0.40 | 2.55 (0.18) |  |
| Expression opinion to friends | 0.32 | 0.17 (0.02) |  |
| Expression opinion to elderly | 0.29 | 0.19 (0.02) |  |
| **Gender Norms** |  |  |  |
| Media use | 0.61 | 1.0 (-) |  |
| Social media use | 0.37 | 0.25 (0.02) |  |
| Having a role model | 0.45 | 0.23 (0.17) |  |
| Being a club member | 0.43 | 0.22 (0.17) |  |
| **Gender Attitudes** |  |  |  |
| Gender Norms Scale F1 | 0.65 | 1.0 (-) |  |
| Gender Norms Scale F2 | 0.55 | 0.79 (0.06) |  |
| Gender Norms Scale F3 | 0.36 | 0.36 (0.03) |  |
| Gender Norms Scale F4 | 0.58 | 0.77 (0.06) |  |
| **Regression Analysis** |  |  |  |
| **Variable** | **$\beta$ (Std. $\beta$)** | **95 % CI** | **p-value** |
| **Outcome: Mental Health** |  |  |  |
| **Gender Attitudes** | -0.50 (-0.57) | [-1.62; 0.62] | p=0.38 |
| **Sex** |  |  |  |
| *Ref.: Male* |  |  |  |
| Female | -0.32 (0.37) | [-1.04; 0.40] | p=0.39 |
| **Gender Roles** | -0.04 (0.08) | [-0.19; 0.11] | p=0.63 |
| **Gender Norms** | 0.50 (0.83) | [-1.13; 2.13] | p=0.55 |
| **Competencies** | **-1.08** (0.40) | [-1.86; -0.30] | p=0.006 |
| **Outcome: Competencies** |  |  |  |
| **Gender Roles** | 0.04 (0.02) | [-0.00; 0.09] | p=0.05 |
| **Gender Norms** | **0.63** (0.07) | [0.49; 0.78] | p<0.001 |
| **Outcome: Gender Attitudes** |  |  |  |
| **Gender Norms** | **1.27** (0.10) | [1.07; 1.48] | p<0.001 |
| **Sex** |  |  |  |
| *Ref.: Male* |  |  |  |
| Female | **0.50** (0.09) | [0.33; 0.67] | p<0.001 |
| **Outcome: Gender Roles** |  |  |  |
| **Gender Norms** | **-1.32** (0.08) | [-1.48; -1.16] | p<0.001 |

Ref. = Reference category; $\beta$ = beta coefficient, Std. $\beta$ = Standardised beta coefficient, CI = Confidence interval, SFL = Standardised factor loading, FL= Factor loading, SE = Standard error, CFI = Bentler Comparative Fit Index, TLI = Tucker-Lewis Index, GFI = Goodness-of-Fit Index, AGFI = Adjusted Goodness-of-Fit Index, RMSEA = Root Mean Square Error of Approximation, SRMR = Standard Root Mean Square Residual, Df = Degrees of freedom
Bold text = $p < 0.05$
Competencies $R^2 = 51.28\%$ ; Gender norms $R^2 = 63.4\%$ ; Gender attitudes $R^2 = 46.6\%$ ; Psychological distress $R^2 = 3.8\%$ ; Gender roles $R^2 = 37.3\%$ ; Sex $R^2 = 7.8\%$
1) The regression analysis was performed simultaneously in one SEM model and was adjusted for: age, work (working in family business or not working in family business), aspiration for higher education (aspiration for secondary school, teacher training, university, or no such aspiration), disability (living with disability or without disability), language spoken in the household (vernacular Afan Oromo, Afar, Amharic or Somali), urbanisation (living in urban or rural area), father living in household (living or not living in household), asset index (sum score capturing material factors in the household), sampling mode (purposeful or non-purposeful), and marital status (married before age 18 or not), regressed on the two exogeneous variables
2) Covariances between: gender norms and gender attitudes, expression of opinion to friends and elderly, gender roles and sex, gender attitudes and psychological distress, having a role model and gender roles based on the modification indices.

**Table 3:** Results of the mediation analysis in the SEM model (N=1,584)

| Variable | Direct effect |  | Indirect effect |  | Total indirect effect |  | Total effect |  | Ratio mediation |
| --- | --- | --- | --- | --- | --- | --- | --- | --- | --- |
| Outcome: Psychological distress |  |  |  |  |  |  |  |  |  |
| β-coefficient (mediator: gender attitudes): |  |  |  |  |  |  |  |  |  |
| Sex | -0.32 | [-1.04; 0.40]<br>p=0.39 | -0.25 | [-0.82; 0.32]<br>p=0.39 | -0.25 | [-0.82; 0.32]<br>p=0.39 | **-0.57** | [-1.00; -0.14]<br>p=0.01 | 44.1% |
| Gender Norms | 0.50 | [-1.13; 2.13]<br>p=0.55 | -0.64 | [-2.05; 0.78]<br>p=0.38 | -0.59 | [-2.08; 0.91]<br>p=0.44 | -0.09 | [-0.56; 0.38]<br>p=0.72 | 734.4% |
| β-coefficient (mediator: gender roles): |  |  |  |  |  |  |  |  |  |
| Gender Norms | 0.50 | [-1.13; 2.13]<br>p=0.55 | 0.05 | [-0.15; 0.25]<br>p=0.63 | -0.59 | [-2.08; 0.91]<br>p=0.44 | -0.09 | [-0.56; 0.38]<br>p=0.72 | 55.8%<sup>N</sup> |
| Outcome: Competencies |  |  |  |  |  |  |  |  |  |
| β-coefficient (mediator: gender roles) |  |  |  |  |  |  |  |  |  |
| Gender Norms | **0.63** | [0.49; 0.78]<br>p<0.001 | -0.06 | [-0.12; 0.001]<br>p=0.055 | -0.06 | [-0.12; 0.001]<br>p=0.055 | **0.57** | [0.47; 0.67]<br>p<0.001 | 10.1 %<sup>N</sup> |

Square brackets = 95% Confidence Intervals Bold text = $p < 0.05$ [] = 95% Confidence interval; p = p-value All values are rounded to the second decimal number <sup>N</sup>=negative ratio

Egalitarian gender attitudes, traditional gender roles, and being a girl (vs. a boy) were also negatively but not significantly associated with psychological distress. In contrast, higher exposure to gender norms was positively and not significantly associated with psychological distress (indicating worse mental health).

We found no statistically significant mediation effects (Table 3). However, the indirect effect of gender norms on competencies via traditional gender roles was close to reaching significance (p=0.055). All estimated indirect effects were negative, except for the pathway from higher exposure to gender norms on psychological distress through traditional gender roles. Notably, the indirect and total indirect effects of higher exposure to gender norms on psychological distress differ, as the former is modelled through two pathways, egalitarian gender attitudes and traditional gender roles, neither of which reached significance. The ratios of the absolute indirect effect to the absolute total effect are 44.1%, 734.4%, 55.8% and 10.1%, respectively.

The unusually large ratio of 734% may result from a total effect close to zero combined with a comparatively large indirect effect estimate. The final three ratios in Table 3 may represent competitive mediations, indicating suppression effects.

## 4 Discussion

### 4.1 Principal Findings

In this study, we empirically tested the hypothesised pathways from the GAM framework (15). We employed SEM to simultaneously model the set of gender-related constructs and their relationships with mental health, including a wide range of covariates, to operationalise the intersectional lens applied in the framework. Based on these data collected from Ethiopian adolescents, five of ten regression paths and none of four mediation paths reached statistical significance.

In the regression models, higher exposure to gender norms was significantly associated with both fewer hours spent on gendered activities (gender roles construct) and more egalitarian gender attitudes. The former association between gender norms (operationalised by being a member of a club, having a role model, as well as media and social media use) and gender roles might be explained by an increased use of (social) media, which was found to be associated with physical inactivity (27). Physical inactivity may also extend to household and caregiving activities. Being enrolled in a club may also contribute to less time spent in the household and, as a result, fewer household and caregiving activities. The association between higher exposure to gender norms and more egalitarian gender attitudes supports a contact hypothesis, arguing that stereotypes are lower when individuals regularly meet, such as in sports clubs (28).

Furthermore, higher exposure to gender norms and traditional gender roles were associated with higher levels of psychological competences, albeit the association for gender roles was not statistically significant. Extracurricular activities have been linked to promoting social and emotional competencies in young people (29). Interactions with other adolescents, such as in a club, can thus contribute to developing adolescents’ competencies and ultimately promote mental health.

Psychological competencies was the only gender-related exposure significantly associated with psychological distress. Thereby, higher levels of competence were negatively associated with psychological distress, indicating better mental health. This is consistent with the literature that psychological competencies are an important determinant of well-being, even in individuals without mental disorders (14).

The associations of egalitarian gender attitudes, traditional gender roles, and being a girl (vs. a boy) with psychological distress were not significant, but their directionalities are still noteworthy. While existing literature indicates that egalitarian gender attitudes are associated with better mental health (12, 30, 14), the other two negative and non-significant associations are somewhat counterintuitive, suggesting that traditional gender roles (i.e., more hours spent on gendered activities) and being an adolescent girl could be protective factors for psychological distress. Findings from countries similar to Ethiopia suggest that adherence to gender roles within a predominantly gendered normative context may contribute to better mental health, as gender roles might be more clearly defined, potentially resulting in less role conflict and a reduced double burden (31). Additionally, violating gender stereotypes in such contexts has been found to produce a backlash and thus negative consequences for mental health (32).

Girls tend to have lower mental health than boys (33). A possible explanation for the non-significant association between sex and psychological distress could be that we controlled for multiple gender-related constructs, which intersect at the individual and meta-level, and without which the negative effects on girls could be reversed. Moreover, the gender gap in mental health was found to be the largest in wealthier and more gender egalitarian countries. In contrast, countries with higher gender inequities exhibit some of the smallest gender gaps in mental health. In some cases, this gap is reversed, with boys reporting worse mental health than girls (31). Additionally, girls in the Ethiopian context might have lower expectations of gender equity, resulting in less feelings of both unmet expectations and relative deprivation (34).

We could not find evidence for mediation effects in our SEM model. However, the directionalities of some non-significant effects are still worth noting. For example, the association between higher exposure to gender norms and psychological distress was positive (i.e., mental health worsens), while more egalitarian gender attitudes negatively mediated this effect, resulting in better mental health. The same directionality is evident in the relationship between gender norms and competencies through gender roles. While higher exposure to gender norms is significantly associated with a higher level of psychological competence, traditional gender roles appear to negatively mediate this effect, resulting in lower levels of psychological competence. These positive direct and negative indirect effects may partly cancel each other out and result in a much smaller coefficient total effect. Although non-significant, this may reflect the complex interplay between the gender-related constructs.

Consequently, the gender-related pathways analysed in the SEM model appear to be more interrelated with each other than directly associated with mental health. Besides the fact that we controlled for gender-related constructs, another possible explanation may be the overall relatively high mental health status of the Ethiopian adolescents. This could have limited the variability in the outcome measure and reduced the ability to detect subtle differences. Moreover, in countries with higher gender inequity, sex differences in mental health tend to be smaller. Traditional gendered expectations may be perceived as normative rather than oppressive (31), thereby mitigating their direct effect on psychological distress. Psychological competencies might be a more proximal factor for psychological distress than the other gender-related constructs and might therefore impact mental health more immediately.

### 4.2 Strengths and Limitations

SEM is an innovative yet still underutilised statistical method for analysing sex/gender in social epidemiology (35, 16). It can be employed to simultaneously investigate potential relationships across multiple variables (16), which is compatible with an intersectional approach (17). We followed reporting guidelines (25) and statistical recommendations for SEM and mediation analysis (e.g., large sample size, the ratio of mediations) (36). We studied indirect and complex relationships through latent variables, which allowed us to model measurement error, as well as multiple regression and mediations (37). Moreover, we used EFA and CFA to ensure the appropriateness of the GAGE gender norms scale for the study population. The findings are representative of Ethiopian adolescents, but may also be relevant for adolescents in countries with similar cultural contexts.

Limitations arise from the GAGE dataset and from our analytical approach. First, we could not examine gender identity due to its absence in the dataset. This is important, as transgender and gender non-conforming adolescents often experience greater discrimination and more pronounced adverse mental health outcomes than their cisgender peers (38). Second, despite being collected by trained interviewers, the data may be subject to social desirability bias. Third, many measures reflect perceived rather than objective measures. To mitigate this limitation, we prioritised measures that were as objective as possible. For example, gender roles were operationalised using time spent on gendered activities rather than attitudes towards gender roles, which are conceptually difficult to distinguish from other gender attitudes.

Limitations also relate to our analytical approach. Some SEM fit statistics fell below recommended thresholds. Several factors may contribute to this, including model complexity, large sample size, numerous covariates necessary to operationalise the intersectional lens, local model misspecifications, and the use of sample weights (37). Since model evaluation should be guided by both theory and statistical fit (37), we assessed model fit by combining theoretical considerations with fit statistics. However, the specified model might not fully capture the underlying data structure. Therefore, findings should be interpreted with caution. Given the complexity of our SEM model, we used cross-sectional data as a first step of theory and plausibility testing. As is common in cross-sectional data, they limit the causal ordering assumption. Future research should seek to use longitudinal data to assess the causality and directionality of the effects.

### 4.3 Conclusions

The gender-related constructs are strongly interlinked, thereby attenuating their individual effects on psychological distress. The interplay of gender-related pathways should be considered when developing interventions to promote mental health in adolescents. Further research should use data that allow for longitudinal study designs to better investigate causality and possible dose-response relationships.

## Data Availability

The data on which the results of our study are based are third-party data that are publicly archived by the data provider UK data service. They are available free of charge following registration with the data provider.

https://www.data-archive.ac.uk/

## 6 Supporting Information A

**S1 Appendix**. Measures.

## 7 Supporting Information B

**S2 Appendix**. Validation of the GAGE gender norms scale for the gender attitudes construct using EFA and CFA.

